# Out-of-Pocket Spending Within 90 Days of Discharge from COVID-19 Hospitalization

**DOI:** 10.1101/2021.06.11.21258766

**Authors:** Kao-Ping Chua, Rena M. Conti, Nora V. Becker

**Affiliations:** Department of Pediatrics, Susan B. Meister Child Health Evaluation and Research Center, University of Michigan Medical School, Ann Arbor; Department of Health Management and Policy, University of Michigan School of Public Health, Ann Arbor; Department of Markets, Public Policy, And Law, Institute for Health System Innovation and Policy, Questrom School of Business, Boston University, Boston; Department of Internal Medicine, Division of General Medicine, University of Michigan Medical School, Ann Arbor

## Abstract

**INTRODUCTION:** Millions of U.S. patients have been hospitalized for COVID-19. After discharge, these patients often have extensive health care needs, but out-of-pocket burden for this care is poorly described. We assessed out-of-pocket spending within 90 days of discharge from COVID-19 hospitalization among privately insured and Medicare Advantage patients.

**METHODS:** In May 2021, we conducted a cross-sectional analysis of the IQVIA PharMetrics^®^ Plus for Academics Database, a national de-identified claims database. Among privately insured and Medicare Advantage patients hospitalized for COVID-19 between March-June 2020, we calculated mean out-of-pocket spending for care within 90 days of discharge. For context, we repeated analyses for patients hospitalized for pneumonia.

**RESULTS:** Among 1,465 COVID-19 patients included, 516 (35.2%) and 949 (64.8%) were covered by private insurance and Medicare Advantage plans. Among these patients, mean (SD) post-discharge out-of-pocket spending was $534 (1,045) and $680 (1,360); spending exceeded $2,000 for 7.0% and 10.3%. Compared with pneumonia patients, mean post-discharge out-of-pocket spending among COVID-19 patients was higher among the privately insured ($534 vs $445) and lower among Medicare Advantage patients ($680 vs $918).

**CONCLUSIONS:** For the privately insured, post-discharge out-of-pocket spending was higher among patients hospitalized for COVID-19 than among patients hospitalized for pneumonia. The opposite was true for Medicare Advantage patients, potentially because insurer cost-sharing waivers for COVID-19 treatment covered the costs of some post-discharge care, such as COVID-19 readmissions. Nonetheless, given the high volume of U.S. COVID-19 hospitalizations to date, our findings suggest a large number of Americans have experienced substantial financial burden for post-discharge care.

## INTRODUCTION

Millions of U.S. patients have been hospitalized for coronavirus disease 2019 (COVID-19).^1^ After discharge, these patients often have extensive health care needs.^2-4^ However, out-of-pocket burden for this care has been poorly described. Addressing this gap in knowledge is crucial, as substantial out-of-pocket burden for post-discharge care might exacerbate other financial stresses experienced by patients hospitalized for COVID-19, such as loss of employment.^4^ Using national claims data from privately insured and Medicare Advantage patients, we assessed out-of-pocket spending within 90 days of discharge from COVID-19 hospitalizations during March-June 2020.

## METHODS

### Study sample

In May 2021, we conducted a cross-sectional analysis of the IQVIA PharMetrics^®^ Plus for Academics Database, which contains medical and pharmacy claims from de-identified patients in all U.S. states. Data included 7.7 million patients covered by fully-insured private plans and 1.0 million patients covered by Medicare Advantage plans in 2020.Claims through September 30, 2020 were available at the time of analysis. Because data were de-identified, the Institutional Review Board of the University of Michigan Medical School exempted analyses from human subjects review.

We identified hospitalizations for privately insured and Medicare Advantage patients that had a primary diagnosis of COVID-19 (ICD-10-CM diagnosis code U071) and that began and ended between March 1-June 30, 2020. We limited analyses to each patient’s first hospitalization during this period. We excluded patients without continuous medical and pharmacy coverage during the 90 days after discharge, patients whose plan was the secondary insurer, and patients with missing out-of-pocket spending data on any claim during the 90 days after discharge.

### Measures

We calculated mean out-of-pocket spending (sum of deductibles, co-insurance, and co-payments) across all claims during the 90 days after discharge. Additionally, we calculated mean out-of-pocket spending for 14 service categories: additional hospitalizations, nursing facility admissions, outpatient (e.g., office visits), emergency department visits, radiology, laboratory, diagnostic/therapeutic procedures (e.g., colonoscopy, surgery), physical/occupational/speech/respiratory therapy, home health/hospice, transportation, clinician-administered medications (e.g., infusions), durable medical equipment and supplies, pharmacy-dispensed prescriptions, and miscellaneous (**Appendix**). For context, we repeated analyses among patients hospitalized for bacterial pneumonia. These patients met similar inclusion and exclusion criterion and did not overlap with the main sample.

### Statistical analyses

Within payer types, we compared post-discharge out-of-pocket spending between COVID-19 and pneumonia patients using a one-part generalized linear model with a log link and Poisson variance function, chosen based on the modified Park test.^5^ Models adjusted for age group, sex, Census region of residence, and month of admission. Analyses used SAS 9.4, Stata 15.1 MP, and two-sided hypothesis tests with α = 0.05.

## RESULTS

Of 2,275 COVID-19 patients meeting inclusion criteria, 810 (35.8%) were excluded, leaving 1,465 patients. **Table 1** displays sample characteristics. Overall, 516 (35.2%) and 949 (64.8%) patients were covered by private insurance and Medicare Advantage plans.

**Table 1.** Characteristics of patients hospitalized for COVID-19 and pneumonia between March-June 2020, IQVIA PharMetrics^®^ Plus for Academics Database

|  | COVID-19 (n= 1,465)<sup>a</sup> |  | Pneumonia (n = 1,374)<sup>b</sup> |  |
| --- | --- | --- | --- | --- |
|  | Private insurance | Medicare Advantage | Private insurance | Medicare Advantage |
| **Number of patients** | 516 | 949 | 245 | 1,129 |
| **Age in years** |  |  |  |  |
| 0-17 | 6 (1.2%) | 0 (0.0%) | 17 (6.9%) | 0 (0.0%) |
| 18-34 | 33 (6.4%) | 4 (0.4%) | 25 (10.2%) | 0 (0.0%) |
| 35-54 | 226 (43.8%) | 44 (4.6%) | 86 (35.1%) | 67 (5.9%) |
| 55-64 | 202 (39.1%) | 102 (10.7%) | 99 (40.4%) | 168 (14.9%) |
| 65-74 | 45 (8.7%) | 319 (33.6%) | 14 (5.7%) | 379 (33.6%) |
| 75-85 | 3 (0.6%) | 319 (33.6%) | 4 (1.6%) | 350 (31.0%) |
| ≥ 86 | 1 (0.2%) | 161 (17.0%) | 0 (0.0%) | 165 (14.6%) |
| **Sex** |  |  |  |  |
| Male | 322 (62.4%) | 411 (43.3%) | 121 (49.4%) | 532 (47.1%) |
| Female | 194 (37.6%) | 538 (56.7%) | 124 (50.6%) | 597 (52.9%) |
| **Region** |  |  |  |  |
| Northeast | 150 (29.1%) | 467 (49.2%) | 25 (10.2%) | 146 (12.9%) |
| Midwest | 122 (23.6%) | 308 (32.5%) | 56 (22.9%) | 617 (54.7%) |
| South | 165 (32.0%) | 110 (11.6%) | 113 (46.1%) | 275 (24.4%) |
| West | 76 (14.7%) | 62 (6.5%) | 51 (20.8%) | 90 (8.0%) |
| **Admission month** |  |  |  |  |
| March | 85 (48.1%) | 143 (54.7%) | 117 (24.1%) | 447 (23.1%) |
| April | 248 (18.8%) | 519 (20.7%) | 59 (16.7%) | 261 (21.8%) |
| May | 97 (16.7%) | 196 (9.6%) | 41 (11.4%) | 246 (15.5%) |
| June | 86 (16.7%) | 91 (9.6%) | 28 (11.4%) | 175 (15.5%) |
| **Mean length of stay (SD)** | 7.9 (8.2) | 8.9 (7.9) | 4.2 (4.2) | 4.6 (3.4) |
| **Any intensive care unit utilization<sup>a</sup>** | 239 (46.3%) | 339 (35.7%) | 86 (35.1%) | 437 (38.7%) |

<sup>a</sup>Among 2,275 patients with an initial hospitalization for COVID-19 between March 1-June 30, 2020, 747 were excluded because they lacked continuous enrollment during the 90 days after discharge, 54 because their insurance was not primary, and 8 because of missing data for out-of-pocket spending. We excluded 1 additional patient owing to the occurrence of a hospitalization that had not resulted in discharge by September 30, 2020 (raising the possibility that out-of-pocket spending for this hospitalization was not fully observed). In total, 810 patients were excluded, leaving 1,465 COVID-19 patients.
<sup>b</sup>We identified 1,810 patients who had a hospitalization with a primary diagnosis code for bacterial pneumonia (ICD-10-CM diagnosis code J13-J18) and no secondary diagnosis code for COVID-19 (U017) between March 1-June 30, 2020, and who were not in the sample of 1,465 COVID-19 patients. Of the 1,813 patients, 365 were excluded because they lacked continuous enrollment during the 90 days after discharge, 47 because their insurance was not primary, 22 because of missing data for out-of-pocket spending, and 2 because they had a hospitalization that had not resulted in discharge by September 30, 2020. In total, 436 patients were excluded, leaving 1,374 pneumonia patients.
<sup>c</sup>Defined as having at least one claim during the hospitalization with a revenue code for intensive care unit (0200-0209) or coronary care unit (0210-0219).

Among privately insured patients, mean (SD) post-discharge out-of-pocket spending was $534 (1,045). For 36 (7.0%) and 15 (2.9%) patients, this spending exceeded $2,000 and $4,000. Service categories accounting for the most out-of-pocket spending were pharmacy-dispensed prescriptions ($130; 24.4% of out-of-pocket spending) and hospitalizations ($111; 20.8%). Among Medicare Advantage patients, mean post-discharge out-of-pocket spending was $680 (1,360). For 98 (10.3%) and 34 (3.6%) patients, this spending exceeded $2,000 and $4,000. Service categories accounting for the most out-of-pocket spending were hospitalizations ($183; 27.0%) and nursing facility admissions ($126; 18.6%) **(Table 2)**. Among privately insured and Medicare Advantage patients, 43.5% and 40.3% of out-of-pocket spending for pharmacy-dispensed prescriptions was for anticoagulants, diabetes drugs (e.g., insulin and sulfonylureas), and bronchodilators. In both populations, these medication classes accounted for the 3 highest shares of out-of-pocket spending for pharmacy-dispensed prescriptions.

**Table 2.**
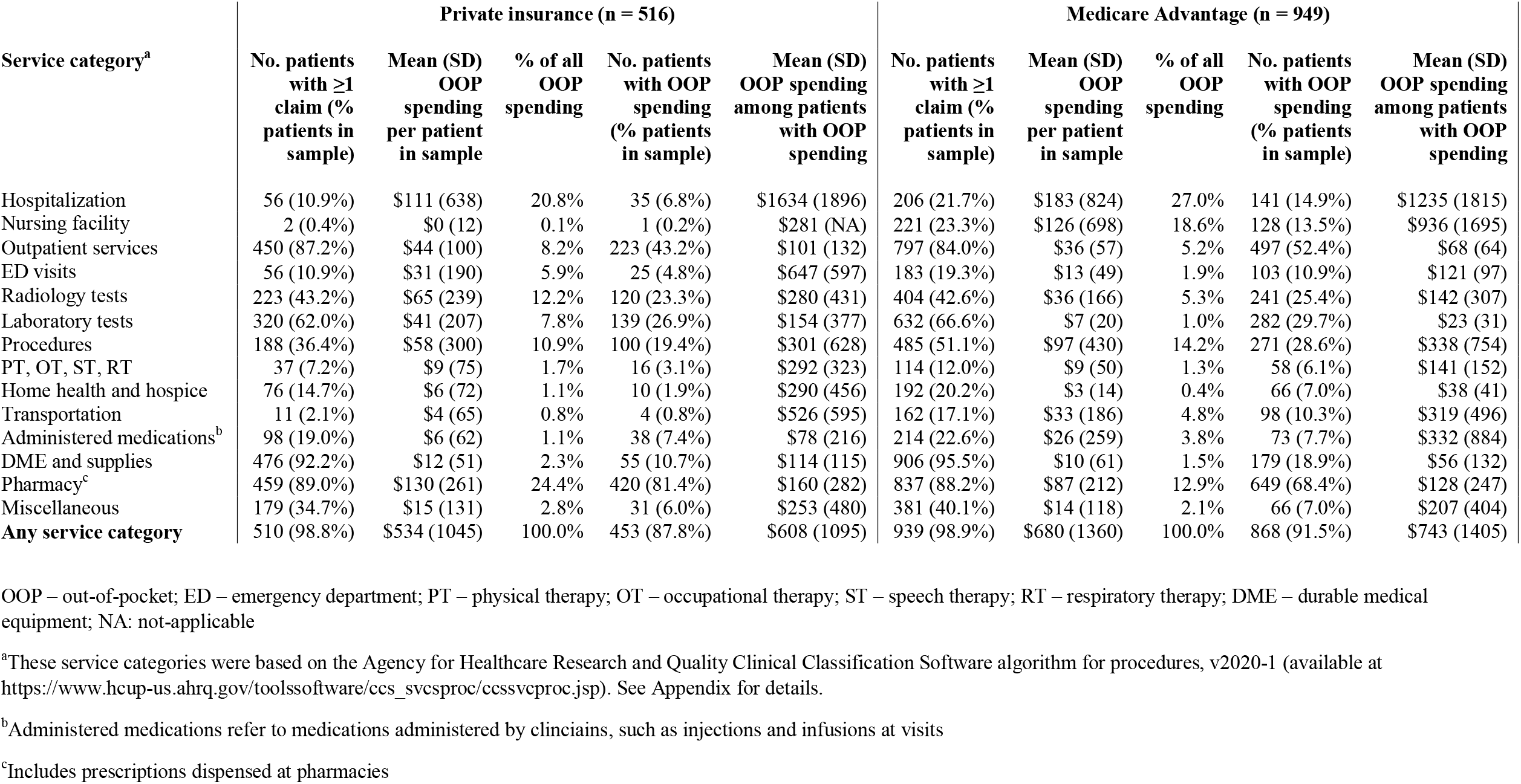
Out-of-pocket spending for health care in the 90 days after COVID-19 hospitalization, IQVIA PharMetrics^®^ Plus for Academics Database

| Service category<sup>a</sup> | Private insurance (n = 516) |  |  |  |  | Medicare Advantage (n = 949) |  |  |  |  |
| --- | --- | --- | --- | --- | --- | --- | --- | --- | --- | --- |
|  | No. patients with ≥1 claim (% patients in sample) | Mean (SD) OOP spending per patient in sample | % of all OOP spending | No. patients with OOP spending (% patients in sample) | Mean (SD) OOP spending among patients with OOP spending | No. patients with ≥1 claim (% patients in sample) | Mean (SD) OOP spending per patient in sample | % of all OOP spending | No. patients with OOP spending (% patients in sample) | Mean (SD) OOP spending among patients with OOP spending |
| Hospitalization | 56 (10.9%) | \$111 (638) | 20.8% | 35 (6.8%) | \$1634 (1896) | 206 (21.7%) | \$183 (824) | 27.0% | 141 (14.9%) | \$1235 (1815) |
| Nursing facility | 2 (0.4%) | \$0 (12) | 0.1% | 1 (0.2%) | \$281 (NA) | 221 (23.3%) | \$126 (698) | 18.6% | 128 (13.5%) | \$936 (1695) |
| Outpatient services | 450 (87.2%) | \$44 (100) | 8.2% | 223 (43.2%) | \$101 (132) | 797 (84.0%) | \$36 (57) | 5.2% | 497 (52.4%) | \$68 (64) |
| ED visits | 56 (10.9%) | \$31 (190) | 5.9% | 25 (4.8%) | \$647 (597) | 183 (19.3%) | \$13 (49) | 1.9% | 103 (10.9%) | \$121 (97) |
| Radiology tests | 223 (43.2%) | \$65 (239) | 12.2% | 120 (23.3%) | \$280 (431) | 404 (42.6%) | \$36 (166) | 5.3% | 241 (25.4%) | \$142 (307) |
| Laboratory tests | 320 (62.0%) | \$41 (207) | 7.8% | 139 (26.9%) | \$154 (377) | 632 (66.6%) | \$7 (20) | 1.0% | 282 (29.7%) | \$23 (31) |
| Procedures | 188 (36.4%) | \$58 (300) | 10.9% | 100 (19.4%) | \$301 (628) | 485 (51.1%) | \$97 (430) | 14.2% | 271 (28.6%) | \$338 (754) |
| PT, OT, ST, RT | 37 (7.2%) | \$9 (75) | 1.7% | 16 (3.1%) | \$292 (323) | 114 (12.0%) | \$9 (50) | 1.3% | 58 (6.1%) | \$141 (152) |
| Home health and hospice | 76 (14.7%) | \$6 (72) | 1.1% | 10 (1.9%) | \$290 (456) | 192 (20.2%) | \$3 (14) | 0.4% | 66 (7.0%) | \$38 (41) |
| Transportation | 11 (2.1%) | \$4 (65) | 0.8% | 4 (0.8%) | \$526 (595) | 162 (17.1%) | \$33 (186) | 4.8% | 98 (10.3%) | \$319 (496) |
| Administered medications<sup>b</sup> | 98 (19.0%) | \$6 (62) | 1.1% | 38 (7.4%) | \$78 (216) | 214 (22.6%) | \$26 (259) | 3.8% | 73 (7.7%) | \$332 (884) |
| DME and supplies | 476 (92.2%) | \$12 (51) | 2.3% | 55 (10.7%) | \$114 (115) | 906 (95.5%) | \$10 (61) | 1.5% | 179 (18.9%) | \$56 (132) |
| Pharmacy<sup>c</sup> | 459 (89.0%) | \$130 (261) | 24.4% | 420 (81.4%) | \$160 (282) | 837 (88.2%) | \$87 (212) | 12.9% | 649 (68.4%) | \$128 (247) |
| Miscellaneous | 179 (34.7%) | \$15 (131) | 2.8% | 31 (6.0%) | \$253 (480) | 381 (40.1%) | \$14 (118) | 2.1% | 66 (7.0%) | \$207 (404) |
| **Any service category** | 510 (98.8%) | \$534 (1045) | 100.0% | 453 (87.8%) | \$608 (1095) | 939 (98.9%) | \$680 (1360) | 100.0% | 868 (91.5%) | \$743 (1405) |

OOP – out-of-pocket; ED – emergency department; PT – physical therapy; OT – occupational therapy; ST – speech therapy; RT – respiratory therapy; DME – durable medical equipment; NA: not-applicable
<sup>a</sup>These service categories were based on the Agency for Healthcare Research and Quality Clinical Classification Software algorithm for procedures, v2020-1 (available at https://www.hcup-us.ahrq.gov/toolssoftware/ccs_svcsproc/ccssvcproc.jsp). See Appendix for details.
<sup>b</sup>Administered medications refer to medications administered by clinicians, such as injections and infusions at visits
<sup>c</sup>Includes prescriptions dispensed at pharmacies

**Table 1** displays characteristics of the 1,374 patients hospitalized for pneumonia included in analyses. Among the 245 privately insured pneumonia patients, mean (SD) post-discharge out-of-pocket spending was $450 (924), compared with $534 among COVID-19 patients (adjusted difference: -$131, 95% CI: -$135, -$128). Among the 1,129 Medicare Advantage patients, mean (SD) post-discharge out-of-pocket spending was $918 (1,263), compared with $680 among COVID-19 patients (adjusted difference: $291, 95% CI: $288, $294). Among 370 Medicare Advantage patients with additional hospitalizations after a pneumonia hospitalization, 306 (82.7%) had out-of-pocket spending for the additional hospitalizations. Among 206 Medicare Advantage patients with additional hospitalizations after a COVID-19 hospitalization, 141 (68.1%) had out-of-pocket spending for the additional hospitalizations.

## DISCUSSION

During the 90 days after COVID-19 hospitalization, privately insured and Medicare Advantage patients paid an average of $534 and $680 for health care; 7.0% and 10.3% of patients paid more than $2,000. For privately insured patients, prescriptions and additional hospitalizations accounted for the highest shares of out-of-pocket spending. For Medicare Advantage patients, additional hospitalizations and nursing facility admissions accounted for the highest shares.

For the privately insured, post-discharge out-of-pocket spending was higher among patients hospitalized for COVID-19 than among patients hospitalized for pneumonia. Perhaps surprisingly, the opposite was true for Medicare Advantage patients. A potential explanation is that some post-discharge care for COVID-19 patients, including readmissions for COVID-19, were covered by insurer cost-sharing waivers for COVID-19 treatment.^6^ In support of this possibility, Medicare Advantage patients who had additional hospitalizations after a COVID-19 hospitalization were less likely to have out-of-pocket spending for the additional hospitalizations compared with pneumonia patients. As many insurers allowed their cost-sharing waivers to expire in early 2021^7^, post-discharge out-of-pocket spending among patients hospitalized for COVID-19 could now be higher than among study patients.

Even if out-of-pocket spending after COVID-19 hospitalization is not greater than for other conditions, our findings suggest a large number of Americans have experienced substantial post-discharge out-of-pocket spending given the volume of COVID-19 hospitalizations to date. To illustrate this point, national data estimate that 2.25 million COVID-19 hospitalizations occurred between August 1, 2020 and June 13, 2021.^1^ If 1 in 14 hospitalizations resulted in post-discharge out-of-pocket spending exceeding $2,000 – the rate for privately insured patients in our study – this would translate to roughly 160,000 hospitalizations and more than $320 million billed to patients for post-discharge care. Importantly, the expiration of insurer cost-sharing waivers means that many patients will now be billed for the COVID-19 hospitalization itself, adding further financial burden beyond that associated with post-discharge care. Consequently, monitoring the financial health of patients hospitalized for COVID-19 is a key policy priority going forward.

This study has limitations. First, analyses only captured the immediate period after COVID-19 hospitalization. Second, the post-discharge period in this study occurred during the early stages of the pandemic, when health care utilization decreased sharply owing partly to social distancing measures.^8^ Post-discharge utilization, and therefore out-of-pocket spending, might be higher now that utilization is rebounding. Third, this study required hospital discharge by June 30, 2020 because a 90-day post-discharge period was needed and claims were complete only through September 30, 2020 at the time of analysis. While necessary, this decision excluded patients with prolonged hospitalizations who may require intensive post-discharge care. Consequently, analyses likely underestimate out-of-pocket burden among all patients hospitalized for COVID-19. Finally, patients may not be fully representative of the entire privately insured and Medicare Advantage population.

## CONCLUSIONS

In this national cohort of 1,465 patients hospitalized for COVID-19 during March-June 2020, out-of-pocket spending within 90 days of discharge exceeded $2,000 for 1 in 14 privately insured patients and 1 in 10 Medicare Advantage patients. Future studies should repeat analyses among patients more recently hospitalized for COVID-19 using longer follow-up periods.

## Data Availability

Data are proprietary and cannot be shared

## Author Contributions

Dr. Chua had full access to all of the data in the study and takes responsibility for the integrity of the data and the accuracy of the data analysis.

Kao-Ping Chua: Conceptualization, Methodology, Software, Formal analysis, Writing – original draft, Data curation, Writing – review and editing, Visualization, Funding acquisition

Rena Conti: Conceptualization, Methodology, Formal analysis, Writing – review and editing

Nora Becker: Conceptualization, Methodology, Formal analysis, Writing – review and editing, Supervision.

## REFERENCES

1. Centers for Disease Control and Prevention. Covid Data Tracker. [Internet]. 2021; https://covid.cdc.gov/covid-data-tracker/#new-hospital-admissions. Accessed June 3, 2021.

2. Lavery AM, Preston LE, Ko JY, et al. Characteristics of Hospitalized Covid-19 Patients Discharged and Experiencing Same-Hospital Readmission - United States, March-August 2020. MMWR Morb Mortal Wkly Rep. 2020;69(45):1695–1699.

3. Weerahandi H, Hochman KA, Simon E, et al. Post-Discharge Health Status and Symptoms in Patients with Severe Covid-19. J Gen Intern Med. 2021;36(3):738–745.

4. Chopra V, Flanders SA, O’Malley M, Malani AN, Prescott HC. Sixty-Day Outcomes among Patients Hospitalized with Covid-19. Ann Intern Med. 2021;174(4):576–578.

5. Buntin MB, Zaslavsky AM. Too Much Ado About Two-Part Models and Transformation? Comparing Methods of Modeling Medicare Expenditures. Journal of Health Economics. 2004;23:525–542.

6. America’s Health Insurance Plans. Health Insurance Providers Respond to Coronavirus (Covid-19). [Internet]. 2021; https://www.ahip.org/health-insurance-providers-respond-to-coronavirus-covid-19. Accessed May 25, 2021.

7. Appleby J. Time to Say Goodbye to Some Insurers’ Waivers for Covid Treatment Fees. [Internet]. 2021; https://khn.org/news/article/time-to-say-goodbye-to-some-insurers-waivers-for-covid-treatment-fees/. Accessed May 25, 2021.

8. Cox C, Amin K, Kamal R. How Have Health Spending and Utilization Changed During the Coronavirus Pandemic? 2021; https://www.healthsystemtracker.org/chart-collection/how-have-healthcare-utilization-and-spending-changed-so-far-during-the-coronavirus-pandemic/#item-start. Accessed June 9, 2021.

