## Appendix for "Out-of-Pocket Spending Within 90 Days of Discharge from COVID-19 Hospitalization"

**Appendix.** Details on methodology

Overview

We identified all medical claims and pharmacy claims during the period spanning the date of discharge to the date of discharge plus 90 days. We excluded any medical claims on the date of discharge associated with the confinement identifier of the index COVID-19 hospitalization. We categorized the remaining claims into 14 service categories. Three of these categories were:

- Hospitalization – this category included any medical claims during the 90-day period associated with a subsequent confinement at a hospital (place of service code 21).
- Nursing facility – this category included any medical claims during the 90-day period associated with a subsequent confinement at a skilled nursing facility (place of service code 31). In our sample, there were no confinements during the 90-day period for confinements at facilities other than hospitals or skilled nursing facilities.
- Pharmacy-dispensed prescriptions – this category included all pharmacy claims for prescriptions dispensed at pharmacies.

The other 11 categories captured all non-confinement-related medical claims during the 90-day post-discharge period: outpatient services, ED visits, radiology, laboratory, procedures, physical therapy/occupational therapy/speech therapy/respiratory therapy, home health and hospice care, transportation, provider-administered medications, durable medical equipment and supplies, and miscellaneous (i.e., all non-confinement-related medical claims not assigned to the first 10 categories). To assign claims to the 11 categories, we used a crosswalk between CPT/HCPCS procedure codes and procedure code categories created by the Agency for Healthcare Research and Quality. This crosswalk, called the Clinical Classification Software for Services and Procedures algorithm (v2020-1), is available for download at this link: <https://www.hcup-us.ahrq.gov/toolssoftware/ccs_svcsproc/ccssvcproc.jsp>

We made minor modifications to the CCS algorithm, such as assigning some codes in CCS category 227 (consultation, evaluation, and preventative care) to different service categories (e.g., assigning office-based evaluation and management codes to the outpatient services category). The table below displays which CCS categories were included in our service categories.

| **Service category** | **CCS Category Number** | **CCS Category description** |
| --- | --- | --- |
| DME and supplies | 241 | Visual aids and other optical supplies |
| DME and supplies | 242 | Hearing devices and audiology supplies |
| DME and supplies | 243 | DME and supplies |
| Emergency department | 227 | Consultation, evaluation, and preventative care^a^ |
| Laboratory | 205 | Arterial blood gases |
| Laboratory | 206 | Microscopic examination (bacterial smear, culture, toxicology) |
| Laboratory | 233 | Laboratory - Chemistry and Hematology |
| Laboratory | 234 | Pathology |
| Laboratory | 235 | Other Laboratory |
| Medications | 222 | Blood and blood product transfusion |
| Medications | 223 | Enteral and parenteral nutrition |
| Medications | 224 | Cancer chemotherapy |
| Medications | 228 | Prophylactic vaccinations and inoculations |
| Medications | 240 | Medications (Injections, infusions and other forms) |
| Miscellaneous | 227 | Consultation, evaluation, and preventative care^a^ |
| Miscellaneous | 237 | Ancillary Services |
| Miscellaneous | 238 | Infertility Services |
| Miscellaneous | 998 | CPTs not classified (F codes) |
| Miscellaneous | 999 | HCPCS Level II codes not classified |
| Home health and hospice | 236 | Nonhospital-based care (e.g., home health care, hospice) |
| Outpatient services | 218 | Psychological and psychiatric evaluation and therapy |
| Outpatient services | 219 | Alcohol and drug management, treatment, and rehabilitation |
| Outpatient services | 220 | Ophthalmologic and otologic diagnosis and treatment |
| Outpatient services | 227 | Consultation, evaluation, and preventative care^a^ |
| Outpatient services | 245 | Telehealth |
| Procedures | 1 | Incision and excision of CNS |
| Procedures | 2 | Insertion, replacement, or removal of extracranial ventricular shunt |
| Procedures | 3 | Laminectomy, excision intervertebral disc |
| Procedures | 4 | Diagnostic spinal tap |
| Procedures | 5 | Insertion of catheter or spinal stimulator and injection into spinal canal |
| Procedures | 6 | Decompression peripheral nerve |
| Procedures | 7 | Other diagnostic nervous system procedures |
| Procedures | 8 | Other non-OR or closed therapeutic nervous system procedures |
| Procedures | 9 | Other OR therapeutic nervous system procedures |
| Procedures | 10 | Thyroidectomy, partial or complete |
| Procedures | 11 | Diagnostic endocrine procedures |
| Procedures | 12 | Other therapeutic endocrine procedures |
| Procedures | 13 | Corneal transplant |
| Procedures | 14 | Glaucoma procedures |
| Procedures | 15 | Lens and cataract procedures |
| Procedures | 16 | Repair of retinal tear, detachment |
| Procedures | 17 | Destruction of lesion of retina and choroid |
| Procedures | 18 | Diagnostic procedures on eye |
| Procedures | 19 | Other therapeutic procedures on eyelids, conjunctiva, cornea |
| Procedures | 20 | Other intraocular therapeutic procedures |
| Procedures | 21 | Other extraocular muscle and orbit therapeutic procedures |
| Procedures | 22 | Tympanoplasty |
| Procedures | 23 | Myringotomy |
| Procedures | 24 | Mastoidectomy |
| Procedures | 25 | Diagnostic procedures on ear |
| Procedures | 26 | Other therapeutic ear procedures |
| Procedures | 27 | Control of epistaxis |
| Procedures | 28 | Plastic procedures on nose |
| Procedures | 29 | Oral and Dental Services |
| Procedures | 30 | Tonsillectomy and/or adenoidectomy |
| Procedures | 31 | Diagnostic procedures on nose, mouth and pharynx |
| Procedures | 32 | Other non-OR therapeutic procedures on nose, mouth and pharynx |
| Procedures | 33 | Other OR therapeutic procedures on nose, mouth and pharynx |
| Procedures | 34 | Tracheostomy, temporary and permanent |
| Procedures | 35 | Tracheoscopy and laryngoscopy with biopsy |
| Procedures | 36 | Lobectomy or pneumonectomy |
| Procedures | 37 | Diagnostic bronchoscopy and biopsy of bronchus |
| Procedures | 38 | Other diagnostic procedures on lung and bronchus |
| Procedures | 39 | Incision of pleura, thoracentesis, chest drainage |
| Procedures | 40 | Other diagnostic procedures of respiratory tract and mediastinum |
| Procedures | 41 | Other non-OR therapeutic procedures on respiratory system |
| Procedures | 42 | Other OR therapeutic procedures on respiratory system |
| Procedures | 43 | Heart valve procedures |
| Procedures | 44 | Coronary artery bypass graft (CABG) |
| Procedures | 45 | Percutaneous transluminal coronary angioplasty (PTCA) |
| Procedures | 46 | Coronary thrombolysis |
| Procedures | 47 | Diagnostic cardiac catheterization, coronary arteriography |
| Procedures | 48 | Insertion, revision, replacement, removal of cardiac pacemaker or cardioverter/defibrillator |
| Procedures | 49 | Other OR heart procedures |
| Procedures | 50 | Extracorporeal circulation auxiliary to open heart procedures |
| Procedures | 51 | Endarterectomy, vessel of head and neck |
| Procedures | 52 | Aortic resection, replacement or anastomosis |
| Procedures | 53 | Varicose vein stripping, lower limb |
| Procedures | 54 | Other vascular catheterization, not heart |
| Procedures | 55 | Peripheral vascular bypass |
| Procedures | 56 | Other vascular bypass and shunt, not heart |
| Procedures | 57 | Creation, revision and removal of arteriovenous fistula or vessel-to-vessel cannula for dialysis |
| Procedures | 58 | Hemodialysis |
| Procedures | 59 | Other OR procedures on vessels of head and neck |
| Procedures | 60 | Embolectomy and endarterectomy of lower limbs |
| Procedures | 61 | Other OR procedures on vessels other than head and neck |
| Procedures | 62 | Other diagnostic cardiovascular procedures |
| Procedures | 63 | Other non-OR therapeutic cardiovascular procedures |
| Procedures | 64 | Bone marrow transplant |
| Procedures | 65 | Bone marrow biopsy |
| Procedures | 66 | Procedures on spleen |
| Procedures | 67 | Other therapeutic procedures, hemic and lymphatic system |
| Procedures | 68 | Injection or ligation of esophageal varices |
| Procedures | 69 | Esophageal dilatation |
| Procedures | 70 | Upper gastrointestinal endoscopy, biopsy |
| Procedures | 71 | Gastrostomy, temporary and permanent |
| Procedures | 72 | Colostomy, temporary and permanent |
| Procedures | 73 | Ileostomy and other enterostomy |
| Procedures | 74 | Gastrectomy, partial and total |
| Procedures | 75 | Small bowel resection |
| Procedures | 76 | Colonoscopy and biopsy |
| Procedures | 77 | Proctoscopy and anorectal biopsy |
| Procedures | 78 | Colorectal resection |
| Procedures | 79 | Local excision of large intestine lesion (not endoscopic) |
| Procedures | 80 | Appendectomy |
| Procedures | 81 | Hemorrhoid procedures |
| Procedures | 82 | Endoscopic retrograde cannulation of pancreas (ERCP) |
| Procedures | 83 | Biopsy of liver |
| Procedures | 84 | Cholecystectomy and common duct exploration |
| Procedures | 85 | Inguinal and femoral hernia repair |
| Procedures | 86 | Other hernia repair |
| Procedures | 87 | Laparoscopy |
| Procedures | 88 | Abdominal paracentesis |
| Procedures | 89 | Exploratory laparotomy |
| Procedures | 90 | Excision, lysis peritoneal adhesions |
| Procedures | 91 | Peritoneal dialysis |
| Procedures | 92 | Other bowel diagnostic procedures |
| Procedures | 93 | Other non-OR upper GI therapeutic procedures |
| Procedures | 94 | Other OR upper GI therapeutic procedures |
| Procedures | 95 | Other non-OR lower GI therapeutic procedures |
| Procedures | 96 | Other OR lower GI therapeutic procedures |
| Procedures | 97 | Other gastrointestinal diagnostic procedures |
| Procedures | 98 | Other non-OR gastrointestinal therapeutic procedures |
| Procedures | 99 | Other OR gastrointestinal therapeutic procedures |
| Procedures | 100 | Endoscopy and endoscopic biopsy of the urinary tract |
| Procedures | 101 | Transurethral excision, drainage, or removal urinary obstruction |
| Procedures | 102 | Ureteral catheterization |
| Procedures | 103 | Nephrotomy and nephrostomy |
| Procedures | 104 | Nephrectomy, partial or complete |
| Procedures | 105 | Kidney transplant |
| Procedures | 106 | Genitourinary incontinence procedures |
| Procedures | 107 | Extracorporeal lithotripsy, urinary |
| Procedures | 108 | Indwelling catheter |
| Procedures | 109 | Procedures on the urethra |
| Procedures | 110 | Other diagnostic procedures of urinary tract |
| Procedures | 111 | Other non-OR therapeutic procedures of urinary tract |
| Procedures | 112 | Other OR therapeutic procedures of urinary tract |
| Procedures | 113 | Transurethral resection of prostate (TURP) |
| Procedures | 114 | Open prostatectomy |
| Procedures | 115 | Circumcision |
| Procedures | 116 | Diagnostic procedures, male genital |
| Procedures | 117 | Other non-OR therapeutic procedures, male genital |
| Procedures | 118 | Other OR therapeutic procedures, male genital |
| Procedures | 119 | Oophorectomy, unilateral and bilateral |
| Procedures | 120 | Other operations on ovary |
| Procedures | 121 | Ligation of fallopian tubes |
| Procedures | 122 | Removal of ectopic pregnancy |
| Procedures | 123 | Other operations on fallopian tubes |
| Procedures | 124 | Hysterectomy, abdominal and vaginal |
| Procedures | 125 | Other excision of cervix and uterus |
| Procedures | 126 | Abortion (termination of pregnancy) |
| Procedures | 127 | Dilatation and curettage (D&C), aspiration after delivery or abortion |
| Procedures | 128 | Diagnostic dilatation and curettage (D&C) |
| Procedures | 129 | Repair of cystocele and rectocele, obliteration of vaginal vault |
| Procedures | 130 | Other diagnostic procedures, female organs |
| Procedures | 131 | Other non-OR therapeutic procedures, female organs |
| Procedures | 132 | Other OR therapeutic procedures, female organs |
| Procedures | 134 | Cesarean section |
| Procedures | 135 | Forceps, vacuum, and breech delivery |
| Procedures | 137 | Other procedures to assist delivery |
| Procedures | 138 | Diagnostic amniocentesis |
| Procedures | 139 | Fetal monitoring |
| Procedures | 140 | Repair of current obstetric laceration |
| Procedures | 141 | Other therapeutic obstetrical procedures including antepartum and postpartum care |
| Procedures | 142 | Partial excision bone |
| Procedures | 143 | Bunionectomy or repair of toe deformities |
| Procedures | 144 | Treatment, facial fracture or dislocation |
| Procedures | 145 | Treatment, fracture or dislocation of radius and ulna |
| Procedures | 146 | Treatment, fracture or dislocation of hip and femur |
| Procedures | 147 | Treatment, fracture or dislocation of lower extremity (other than hip or femur) |
| Procedures | 148 | Other fracture and dislocation procedure |
| Procedures | 149 | Arthroscopy |
| Procedures | 150 | Division of joint capsule, ligament or cartilage |
| Procedures | 151 | Excision of semilunar cartilage of knee |
| Procedures | 152 | Arthroplasty knee |
| Procedures | 153 | Hip replacement, total and partial |
| Procedures | 154 | Arthroplasty other than hip or knee |
| Procedures | 155 | Arthrocentesis |
| Procedures | 156 | Injections and aspirations of muscles, tendons, bursa, joints and soft tissue |
| Procedures | 157 | Amputation of lower extremity |
| Procedures | 158 | Spinal fusion |
| Procedures | 159 | Other diagnostic procedures on musculoskeletal system |
| Procedures | 160 | Other therapeutic procedures on muscles and tendons |
| Procedures | 161 | Other OR therapeutic procedures on bone |
| Procedures | 162 | Other OR therapeutic procedures on joints |
| Procedures | 163 | Other non-OR therapeutic procedures on musculoskeletal system |
| Procedures | 164 | Other OR therapeutic procedures on musculoskeletal system |
| Procedures | 165 | Breast biopsy and other diagnostic procedures on breast |
| Procedures | 166 | Lumpectomy, quadrantectomy of breast |
| Procedures | 167 | Mastectomy |
| Procedures | 168 | Incision and drainage, skin and subcutaneous tissue |
| Procedures | 169 | Debridement of wound, infection or burn |
| Procedures | 170 | Excision of skin lesion |
| Procedures | 171 | Suture of skin and subcutaneous tissue |
| Procedures | 172 | Skin graft |
| Procedures | 173 | Other diagnostic procedures on skin and subcutaneous tissue |
| Procedures | 174 | Other non-OR therapeutic procedures on skin and breast |
| Procedures | 175 | Other OR therapeutic procedures on skin and breast |
| Procedures | 176 | Other organ transplantation |
| Procedures | 199 | Electroencephalogram (EEG) |
| Procedures | 200 | Nonoperative urinary system measurements |
| Procedures | 201 | Cardiac stress tests |
| Procedures | 202 | Electrocardiogram |
| Procedures | 203 | Electrographic cardiac monitoring |
| Procedures | 204 | Swan-Ganz catheterization for monitoring |
| Procedures | 214 | Traction, splints, and other wound care |
| Procedures | 216 | Respiratory intubation and mechanical ventilation |
| Procedures | 221 | Nasogastric tube |
| Procedures | 225 | Conversion of cardiac rhythm |
| Procedures | 229 | Nonoperative removal of foreign body |
| Procedures | 230 | Extracorporeal shock wave, other than urinary |
| Procedures | 231 | Other therapeutic procedures |
| Procedures | 232 | Anesthesia |
| Procedures | 244 | Gastric bypass and volume reduction |
| PT/OT/ST/RT | 212 | Diagnostic physical, occupational, and speech therapy |
| PT/OT/ST/RT | 213 | Physical, occupational, and speech therapy exercises; manipulation; and other procedures |
| PT/OT/ST/RT | 215 | Other physical, occupational, and speech therapy and rehabilitation |
| PT/OT/ST/RT | 217 | Other respiratory therapy |
| Radiology | 177 | Computerized axial tomography (CT) scan head |
| Radiology | 178 | CT scan chest |
| Radiology | 179 | CT scan abdomen |
| Radiology | 180 | Other CT scan |
| Radiology | 181 | Myelogram |
| Radiology | 182 | Mammography |
| Radiology | 183 | Routine chest X-ray |
| Radiology | 184 | Intraoperative cholangiogram |
| Radiology | 185 | Upper gastrointestinal X-ray |
| Radiology | 186 | Lower gastrointestinal X-ray |
| Radiology | 187 | Intravenous pyelogram |
| Radiology | 189 | Contrast aortogram |
| Radiology | 190 | Contrast arteriogram of femoral and lower extremity arteries |
| Radiology | 191 | Arterio- or venogram (not heart and head) |
| Radiology | 192 | Diagnostic ultrasound of head and neck |
| Radiology | 193 | Diagnostic ultrasound of heart (echocardiogram) |
| Radiology | 194 | Diagnostic ultrasound of gastrointestinal tract |
| Radiology | 195 | Diagnostic ultrasound of urinary tract |
| Radiology | 196 | Diagnostic ultrasound of abdomen or retroperitoneum |
| Radiology | 197 | Other diagnostic ultrasound |
| Radiology | 198 | Magnetic resonance imaging |
| Radiology | 207 | Radioisotope bone scan |
| Radiology | 208 | Radioisotope pulmonary scan |
| Radiology | 209 | Radioisotope scan and function studies |
| Radiology | 210 | Other radioisotope scan |
| Radiology | 211 | Therapeutic radiology |
| Radiology | 226 | Other diagnostic radiology and related techniques |
| Transportation | 239 | Transportation - patient, provider, equipment |

^a^Procedure codes in CCS category 227 specific to the emergency department were assigned to “emergency department”; those for outpatient services were assigned to “outpatient services”, and the remainder were assigned to “miscellaneous”
